# What Does an Agent-Run Clinic Cost? Protocol for a Prospective Observational Study of the Compute Cost of Completed Work in a Clinical Agent Fleet

**DOI:** 10.64898/2026.11.19.26368538

**Authors:** Sofian Youssef, Omar Elalfy, Joseph Davids

## Abstract

**Background:** Decisions about deploying large language model (LLM) agents in health care require the cost of completed work in production. Published evidence reports benchmark accuracy, benchmark-level cost, and time or financial outcomes for ambient documentation, and list prices alone do not determine the cost of completed work. To our knowledge, no deployed clinical agent fleet has published a measurement method that ties compute cost, latency, failures and retries to completed work.

**Objective:** This study aims to estimate the compute cost of completed work by the orchestrated clinical agents of a deployed LLM agent fleet over a 12-week window of production telemetry, using estimators, data-quality gates and an analysis plan fixed before any window data are extracted for the study.

**Methods:** The setting is a private outpatient clinic in London, United Kingdom, where wai, a multi-agent system supporting clinic operations, is deployed. The population is every run of the 20 orchestrated clinical documentation and decision-support agents starting between September 21 and December 13, 2026. The run store writes 2 records for every run, a non-terminal record before the model call and a terminal record after it, and has no attempt counter or parent-run link. Estimators defined on that structure cover cost per resolved execution and per completed intention, retry and failed-intention shares of compute, error and failure rates, latency, cost per draft offered for clinician review and a latency-cost frontier, by agent, task class and acceptance stratum. Cost is recorded in GBP pence, converted from provider-returned US dollar cost at a fixed rate and rounded per model call. The plan fixes weekly aggregation, small-cell suppression, bootstrap intervals over weeks, 2 prespecified comparisons, 5 sensitivity analyses and 4 data-quality gates whose failure converts an estimate into a bound or an unmeasured stratum. Two staff-level measures proposed during planning were dropped. Revenue, pricing, margin and staffing inference are excluded.

**Results:** The measurement window opened on September 21, 2026, and closes on December 13, 2026, and telemetry is being recorded in routine operation. No window data had been extracted for this study at submission. Aggregate whole-table counts that include window records were extracted on November 19, 2026, for companion manuscripts and are disclosed. Extraction will begin no earlier than December 15, 2026, and the results are expected to be submitted for publication in 2027.

**Conclusions:** The compute cost of completed clinical agent work can be specified as estimators over the run-level telemetry this deployment records, provided paired recording rows are separated from retries, missing terminal states are bounded and rounding of recorded cost is reported. This protocol fixes those estimators, and the reporting consequence of each known instrumentation defect, before extraction.

**Trial Registration:** None. The analysis plan will be registered on Open Science Framework Registries before any window data are extracted for this study.

International Registered Report Identifier (IRRID): DERR1-10.2196/ (to be assigned by the journal)

## Introduction

### Background

Health systems are moving large language model (LLM) tools from drafting text toward agents that take actions in clinical operations. Ambient documentation is, to our knowledge, the clinical LLM application with the most mature evidence, including randomized trials. Two pragmatic trials published in December 2025 found small or inconsistent reductions in documentation time, with a significant reduction in work exhaustion in one trial and a direction of benefit that requires confirmation in the other [1,2]. The accompanying editorial anticipated that next-generation products would extend into orders, prior authorization and billing [3]. A 5-site difference-in-differences study of 8581 clinicians estimated 16.0 fewer documentation minutes per 8 scheduled hours among adopters [4]. Agent research has advanced in parallel. Agents have been benchmarked in a virtual electronic health record (EHR) environment [5] and evaluated in a sandboxed EHR able to order tests and prescribe [6], and a prospective deployment of an agentic workflow for hospital course summaries reported its safety outcomes [7]. An evidence map found early agent deployments concentrated in administrative workflows [8]. A systematic review of 519 LLM evaluations found that only 5% used real patient care data [9].

The economic evidence for these systems is thinner than the evidence on accuracy and time. For ambient scribes, a single-system analysis of more than 1.2 million ambulatory encounters reported changes in physician financial productivity [10], and an invited commentary set these gains against per-clinician subscription fees [11]. A national evaluation of ambient voice technology in the National Health Service (NHS) in England that includes cost-consequence and budget-impact modeling is at the protocol stage [12]. These analyses price a tool by subscription and measure outcomes at the level of the clinician or the encounter. They do not expose the compute cost of a unit of work. Benchmark studies have begun to report computational cost alongside accuracy [13], to show that concatenating clinical tasks reduces cost under controlled load [14], and to relate accuracy to token budgets across model sizes [15]. Outside medicine, an analysis of agent benchmarks argued that agents should be evaluated jointly on cost and accuracy [16]. Two 2026 preprints report figures close to production. The governance framework for an EHR-embedded documentation agent tracks cost and latency and reports completion after retries absorbed transient model errors [17], and a simulated scheduling agent reports median latency and an average operational cost per appointment [18]. A study of 983,154 patient message responses by 1542 clinicians found that adoption of a drafting tool was associated with longer message turnaround, although use of a draft was associated with shorter turnaround, which indicates that latency is an operational outcome as well as an engineering metric [19].

Trends in inference prices complicate the measurement problem. The price of reaching a fixed level of benchmark performance fell by between about 9-fold and 900-fold per year depending on the performance milestone [20]. A subsequent analysis of the largest price dataset assembled to date estimated that the price of a given level of performance fell about 5-fold to 10-fold per year for frontier models, while the price of running frontier models rose between 3-fold and 18-fold per year because of larger models and greater reasoning demands [21]. The cost of completed work therefore depends on which model is called, how many tokens each task consumes, how calls are orchestrated, and how often they fail or are retried, and the list price sets none of these.

The economically relevant unit is a completed piece of work rather than a single model generation, because runs fail, are retried and fan out across agents. Cost per generation omits the compute spent on failed and retried attempts. Computing the cost of completed work requires that every run is recorded, that a retry can be distinguished from a second record of the same run, and that missing terminal states can be detected. Regulators have asked for related logging for other purposes. NHS England guidance for ambient scribing expects deployer monitoring independent of the manufacturer [22], the United Kingdom National Commission into the Regulation of AI in Healthcare recommended logging to detect performance drift [23], and the European Union Artificial Intelligence Act requires automatic event logging and log retention by providers and deployers of high-risk systems [24]. The Medicines and Healthcare products Regulatory Agency (MHRA) AI Airlock report found that human oversight may not function as a fixed safety control over a device’s lifecycle [25]. Reporting standards for health economic evaluation, including the extension for interventions that use artificial intelligence, require transparent reporting of resource use [26,27], and reporting guidance exists for studies that use LLMs [28] and for analyses of EHR-use metadata [29]. To our knowledge, no deployed clinical agent fleet has published a measurement framework that links compute cost, latency, failures and retries to completed work.

### Objective

This study aims to estimate the compute cost of completed work by the orchestrated clinical agents of an LLM agent fleet over a prespecified 12-week window of production telemetry. This protocol sets out the units of analysis, the estimators, the rules for separating recording rows from retries, the analysis plan with its denominators and failure conditions, and the state of the deployment’s instrumentation before the window, so that each is fixed and publicly registered before any window data are extracted for the study.

## Methods

### Study Design

This is a single-site prospective observational study of routinely recorded production telemetry over a prespecified 12-week window, with an analysis plan registered before extraction. The study is not an economic evaluation. It measures compute resource use per unit of work, which could serve as an input to an evaluation reported under the Consolidated Health Economic Evaluation Reporting Standards 2022 (CHEERS 2022) and its extension for artificial intelligence (CHEERS-AI) [26,27]. Reporting follows the applicable items of TRIPOD-LLM [28], with items that do not apply to a protocol marked as such (Multimedia Appendix 2).

### Setting and System

The setting is a private outpatient clinic in London, United Kingdom. The system is wai, a multi-agent system supporting clinic operations, developed and operated by Wai Health Inc, which supports patient messaging, voice contact, scheduling, documentation drafting, results handling and proactive monitoring. Its architecture separates proposal from commitment: agents propose actions, and for actions that cross the approval boundary a person accepts the proposal before the effect is committed [30]. Version 0.2 of the code-derived action inventory (September 8, 2026) records 153 agent actions in 23 functional domains [30,31]. The deployment is described in a companion manuscript [32].

This study covers the orchestrated clinical pipeline, the part of the fleet whose every run is written to a single run store. The pipeline comprises 20 agents that draft clinical summaries, patient summaries, letters, sick notes and clinical notes, extract action points, and return decision support to clinicians (risk screening, differential diagnosis, safetynetting,drug interaction checking, coding, guideline lookup, investigation suggestion, risk scores, mental health risk triage, medication review, fit-to-fly assessment and note critique). They correspond to rows AA-01 to AA-20 of the inventory. Other agents in the fleet, including the messaging, voice and operations agents, do not write to the run store, and the call-level model log that some of them write carries no run identifier, so their compute cannot be attributed to units of work and is outside this study. As an indication of scale, the run store held 13,250 records written by 19 agents between May 24 and September 20, 2026 (6643 non-terminal, 5880 done and 727 error), counted under a version-locked extraction of pre-window records only.

Models The orchestrator assigns each agent a model tier, and each tier resolves through a configured chain of models accessed through OpenRouter. In the production configuration read on November 19, 2026, the standard, low-cost and critic tiers used by the 20 agents resolved to deepseek/deepseek-v4-pro (DeepSeek), with deepseek/deepseek-v4-pro and then z-ai/glm-5.2 (Z.ai) as fallbacks, and 2 guidance skills named deepseek/deepseek-v4-flash directly. The date on which each live configuration value was set is not recorded, and the code changes affecting model selection that were committed during the window are listed in Multimedia Appendix 1. The model mix during the window is therefore measured from the data rather than taken from configuration. The terminal record of each run stores the model that answered, whereas the non-terminal record stores the model requested, so the model mix is computed from terminal records only. Training cutoff dates of the provider models are not available to the authors. Prompt versions are recorded per run.

### Data Sources

The primary data source is the run store, which records each orchestrated run with the agent identifier, model identifier, input hash, output hash, input and output token counts, latency, cost, prompt version, status, start time and finish time. Status takes one of 6 values: queued, running, done, error, skipped or cost-capped. The run store holds no direct patient identifier. Each record carries an encounter identifier and a free-text error field, neither of which is extracted. The secondary sources are the action inventory version 0.2 [30,31], which maps each agent to its acceptance class, and the orchestrator code and configuration at commit 687f45e295, which define how records, cost and retries are written. The call-level model log, the communications approval record and the audit trail are not used.

### How a Run Is Recorded

Every orchestrated execution writes 2 records. The orchestrator inserts a non-terminal record with status running before the model call and then inserts a second, separate record with the terminal status, the token counts, cost, latency and output hash after it. The first record is never updated or removed. On the normal path, usage, cost and terminal status are written together in the second insert, so if that insert is lost the evidence that the run consumed tokens is lost with it. When the model call or its retry raises an exception, the error record is written with no token counts, cost or latency, even if an earlier attempt had consumed tokens.

Retries occur at 3 levels, and none is linked in the run store. Within a call, a failed model candidate fails over to the next model in the tier’s chain, and the failed attempt’s usage is not recorded. Within a run, an output that fails parsing, schema validation or the output-language check is retried once, preferring a different model, and the tokens, cost and latency of both attempts are summed into the one terminal record. Across runs, a failed job is requeued with exponential backoff for up to 3 attempts, and a job whose worker died is requeued after a stale period. The run store has no attempt counter, parent-run identifier or job identifier. A requeued job produces the same input hash only if its payload and idempotency key are unchanged, and one document path includes the current time in its key, so its retries always appear as new intentions.

Recorded cost is in GBP pence, although the field name refers to cents. For each model call, the orchestrator takes the cost returned by OpenRouter when the response carries one and converts it at a fixed 79 pence per US dollar, rounding to a whole penny per call. Only when no cost is returned does it multiply token usage by a rate table held in code, last changed on July 14, 2026, which assigns zero cost to any model it does not list. Per-call values are summed over tool turns and the schema retry into the run’s cost. Because each call is rounded to a whole penny, many low-cost runs record 0: 31.3% of done records before the window recorded a cost of 0. Cache tokens are priced but not stored, so recorded token counts cannot reproduce recorded cost.

### Units of Analysis

The units of analysis are defined in Table 1, and the measurement chain is shown in Figure 1. The central distinction is between a record, an execution and an intention. A record is a row in the run store. An execution is 1 orchestrated attempt, represented by its terminal record, or by an unpaired non-terminal record when the terminal record is missing. An intention is the unit of work an agent was asked to perform, which may require more than 1 execution if earlier attempts fail. Executions are grouped into intentions by the intention key, the agent identifier together with the input hash, so that 2 agents given the same input are never merged. Grouping by key identifies only retries across runs that reuse the same input. Failover within a call and the schema retry within a run are inside 1 execution, and retries that change their input appear as new intentions, which biases the retry share downward and intention failure upward.

**Table 1.** Units of analysis and definitions used by the framework.

| Unit | Definition | Use |
|---|---|---|
| Record | A row in the run store | Raw input to the recording rules |
| Recording row | A non-terminal record with no token usage or cost that shares agent identifier, input hash and start time with a terminal record, and is paired 1 to 1 with it (rule D1) | Excluded from all numerators and denominators |
| Execution | A terminal record, or a non-terminal record left unpaired by rule D1 | Unit of compute cost and of run-level rates |
| Resolved execution | An execution with a terminal status (done, error, skipped or cost-capped) | Denominator for cost per resolved execution |
| Completed execution | An execution with terminal status done | Run-level completion |
| Intention key | Agent identifier together with input hash | Grouping key for intentions |
| Intention | The executions sharing an intention key that started within 24 hours of the first execution in the group (rule D4) | Unit of work for retry and completion estimators |
| Completed intention | An intention with at least 1 completed execution | Denominator for cost per completed intention |
| Acceptance stratum | Grouping of agents by the acceptance class of their inventory row: review required, acceptance not enforced on every path, or advisory | Stratification and estimator E6 |
| Task class | Prespecified grouping of agents by workload: fast triage, deep reasoning or other | Stratification and prespecified comparisons |
| Configuration | A combination of agent, model identifier on the terminal record and prompt version | Unit of the latency-cost frontier |

The fast triage class comprises the red-flag, mental health risk triage, safety-netting, drug interaction and action point agents. The deep reasoning class comprises the note composition, investigation suggestion and guideline lookup agents. The remaining 12 agents form the third class. The acceptance strata follow the inventory's own accept column. The review-required stratum comprises the 7 agents whose output is a draft that a clinician must accept or sign before it takes effect (clinical summary, patient summary, general practitioner letter, referral letter, sick note, letter composer and note composer). The stratum without enforced acceptance comprises the 2 agents classed Partial, which accept on some code paths and not on others (action points and fit-to-fly assessment); they are assigned there as the conservative choice. The advisory stratum comprises the 11 agents whose output is shown to a clinician and enters no record or patient channel unaided. The complete agent map, with inventory row numbers, is frozen in Multimedia Appendix 1. Any agent identifier that appears in the window and is not in the map is assigned to the other task class and mapped to its inventory row by module path before extraction of estimates, or reported as unmapped.

**Figure 1.**
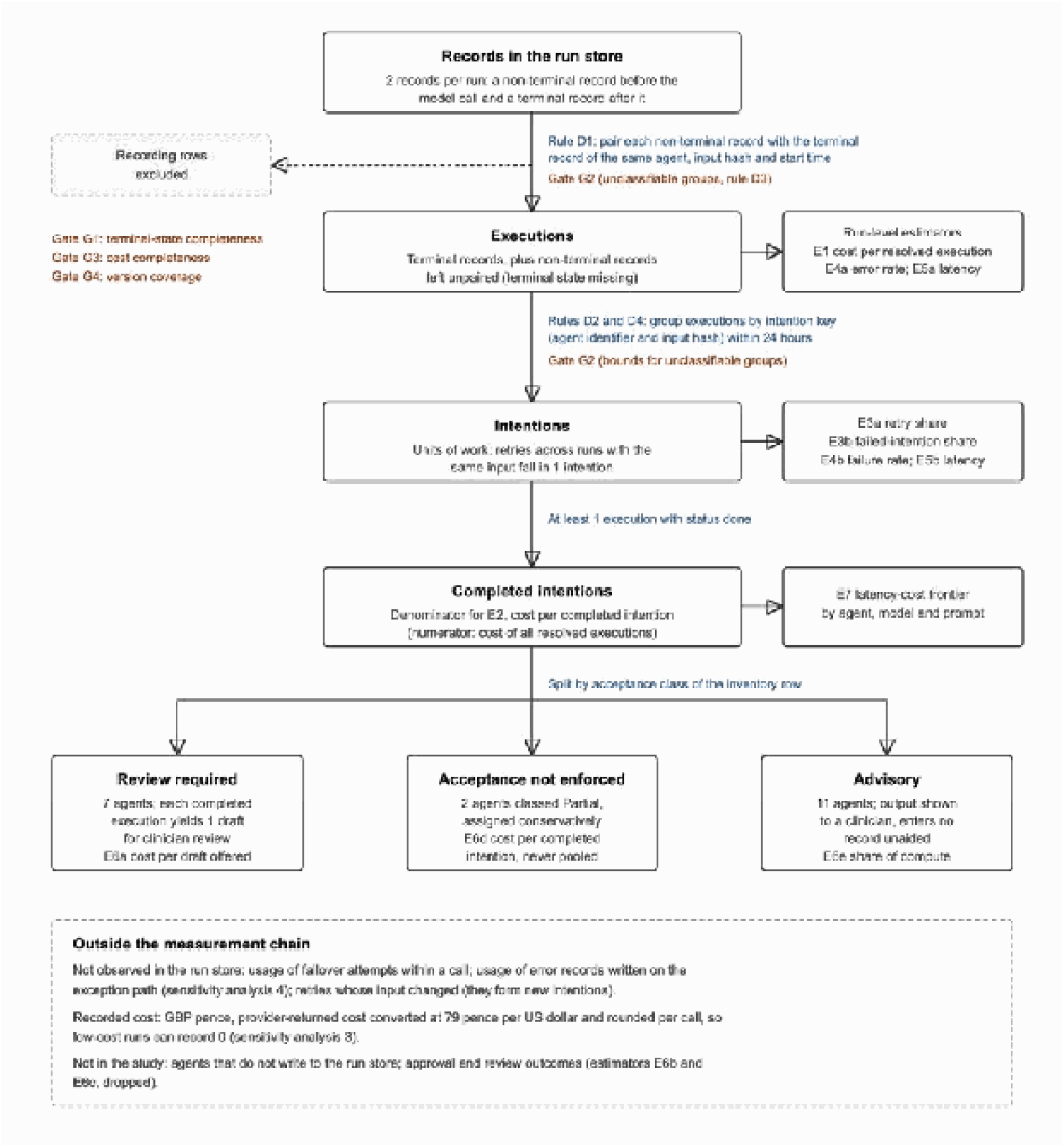
Measurement chain from records in the run store to completed intentions and acceptance strata. Recording rows are removed by pairing (rule D1), executions are grouped into intentions (rules D2 to D4), and completed intentions are split by the acceptance class of each agent’s inventory row. Each transition is annotated with the rule, the estimators it feeds (E1 to E7) and the data-quality gates that govern it (G1 to G4). The lower panel lists what the run store does not observe and what is outside the study.

### Estimators

The estimators are summarized in Table 2. All cost estimators are ratios of sums, so that the reported unit cost multiplied by the reported count reproduces the stratum’s recorded compute cost. No total expenditure is reported. Cost per execution is written c and is the recorded cost in GBP pence.

**Table 2.**
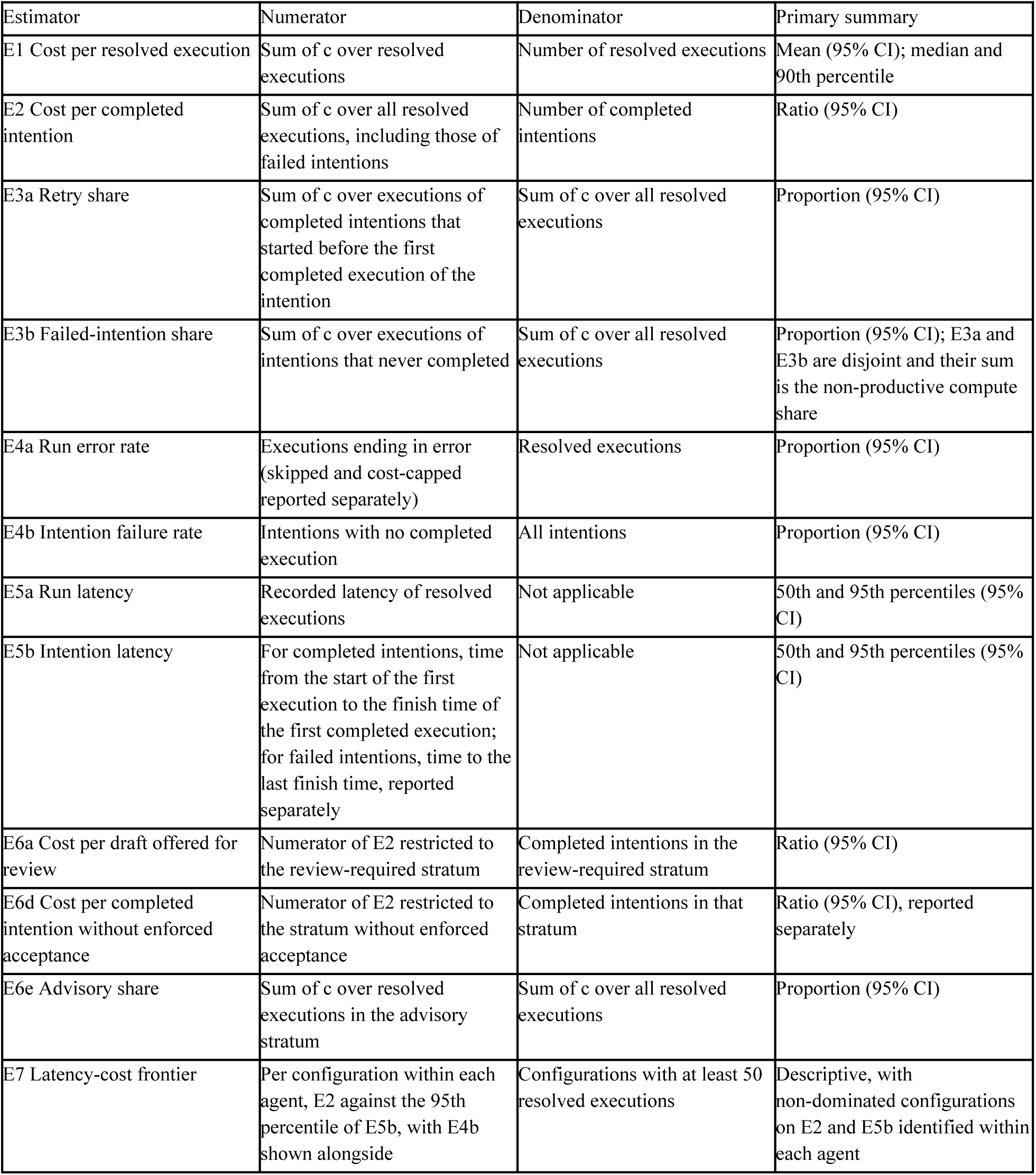
Prespecified estimators, numerators and denominators.

| Estimator | Numerator | Denominator | Primary summary |
|---|---|---|---|
| E1 Cost per resolved execution | Sum of c over resolved executions | Number of resolved executions | Mean (95% CI); median and 90th percentile |
| E2 Cost per completed intention | Sum of c over all resolved executions, including those of failed intentions | Number of completed intentions | Ratio (95% CI) |
| E3a Retry share | Sum of c over executions of completed intentions that started before the first completed execution of the intention | Sum of c over all resolved executions | Proportion (95% CI) |
| E3b Failed-intention share | Sum of c over executions of intentions that never completed | Sum of c over all resolved executions | Proportion (95% CI); E3a and E3b are disjoint and their sum is the non-productive compute share |
| E4a Run error rate | Executions ending in error (skipped and cost-capped reported separately) | Resolved executions | Proportion (95% CI) |
| E4b Intention failure rate | Intentions with no completed execution | All intentions | Proportion (95% CI) |
| E5a Run latency | Recorded latency of resolved executions | Not applicable | 50th and 95th percentiles (95% CI) |
| E5b Intention latency | For completed intentions, time from the start of the first execution to the finish time of the first completed execution; for failed intentions, time to the last finish time, reported separately | Not applicable | 50th and 95th percentiles (95% CI) |
| E6a Cost per draft offered for review | Numerator of E2 restricted to the review-required stratum | Completed intentions in the review-required stratum | Ratio (95% CI) |
| E6d Cost per completed intention without enforced acceptance | Numerator of E2 restricted to the stratum without enforced acceptance | Completed intentions in that stratum | Ratio (95% CI), reported separately |
| E6e Advisory share | Sum of c over resolved executions in the advisory stratum | Sum of c over all resolved executions | Proportion (95% CI) |
| E7 Latency-cost frontier | Per configuration within each agent, E2 against the 95th percentile of E5b, with E4b shown alongside | Configurations with at least 50 resolved executions | Descriptive, with non-dominated configurations on E2 and E5b identified within each agent |

Three features of these definitions follow from the measurement problem. First, E2 places the cost of failed and retried executions in the numerator and only completed intentions in the denominator. Because E1 and E2 share a numerator, the ratio of E2 to E1 equals the number of resolved executions per completed intention. It measures the volume of failure and retry, and the non-productive compute share (E3a plus E3b) gives the cost-weighted counterpart. Second, each completed execution of a review-required agent persists 1 draft for clinician review, so E6a is the compute cost of a draft offered for review, including the compute spent on failed attempts. It is not a cost per accepted draft. Third, agents without enforced acceptance are never pooled with review-required agents, because pooling would lower the apparent cost of reviewed work by including actions that bypass review.

The plan initially included 2 further estimators, review yield (E6b) and cost per committed effect (E6c), which aggregate staff review decisions; both were dropped because they would require analysis of staff-level approval data, which this study does not use. With them, the proposal-to-effect lineage and its unlinked-share residual (E6f) were removed, because no linkage beyond the run store is required.

### Recording, Duplicate and Retry Rules

The rules are defined on the 2-record structure described above. In a version-locked extraction of pre-window records (May 24 to September 20, 2026), 6605 of 6643 non-terminal records shared agent identifier, input hash and exactly the same start time with a terminal record, and none carried token usage or cost. Of 6604 intention-key groups, 6537 (99.0%) were exactly 1 non-terminal and 1 terminal record, 31 were a single non-terminal record with no terminal record, and 36 were larger groups. No key had 2 done records. The framework applies 4 prespecified rules.

- Rule D1. A non-terminal record with no token usage or cost that shares agent identifier, input hash and start time with a terminal record is a recording row and is excluded. Pairing is 1 to 1 in start-time order, so that a group of 2 non-terminal and 2 terminal records yields 2 executions.
- Rule D2. Terminal records sharing an intention key are separate executions of 1 intention, and all of their costs are counted. A non-terminal record left unpaired by rule D1 is an execution whose terminal state is missing and is handled under gate G1.
- Rule D3. Groups that rule D1 cannot pair 1 to 1, for example 2 non-terminal records with the same start time and 1 terminal record, are unclassifiable. For these groups, intentionlevelestimators are computed under a lower bound, in which every unpaired non-terminal record is treated as a recording row, and an upper bound, in which every unpaired non-terminal record is treated as a failed execution.
- Rule D4. Executions sharing an intention key form 1 intention only if they started within 24 hours of the first execution in the group. Later executions with the same key start a new intention, which accommodates scheduled re-evaluation of unchanged inputs. Grouping windows of 1 hour and 7 days are sensitivity analyses. A unique index on encounter, agent and input hash applies to done records only. It prevents 2 completed records of the same work but does not constrain non-terminal or error records. Neither a fix for the 2-record write path nor a process that resolves non-terminal records in the run store existed at commit 687f45e295 (November 19, 2026). If either is deployed during the window, its first effect visible in the run store is treated as a change point and estimators are reported for the periods before and after it.

### Prespecified Analysis Plan

The measurement window is 12 consecutive ISO weeks, weeks 39 to 50 of 2026, from Monday, September 21, 2026, 00:00 to Sunday, December 13, 2026, 23:59 London time. All executions that started within the window are eligible, and telemetry is aggregated by ISO week in London time. Runs that started within the window are followed for 24 hours after the window closes to allow terminal records to be written. The run store has no field that marks a test or sandbox run, so runs generated by staff testing in the production environment cannot be separated and are included.

Estimates are reported for the whole pipeline, for each task class, for each acceptance stratum, for each agent, and by configuration for the latency-cost frontier. Weekly cells with fewer than 5 events are suppressed in any published table. An estimator whose denominator is below 50 in a stratum over the window (resolved executions for E1 to E5, completed intentions for E6a and E6d) is reported as unmeasured for that stratum and is not pooled with neighboring strata.

Cost estimators are ratios of sums. Proportions are reported with their numerators and denominators. Intervals are 95% percentile intervals from a nonparametric bootstrap that resamples ISO weeks with replacement (2000 replicates), which preserves within-week correlation between runs. Because 12 weeks give limited information about between-week variance, and percentile intervals from few clusters tend to undercover, these intervals will be interpreted as approximate. The weekly series is the primary display, and the number of weeks contributing to each interval is reported. Latency quantiles are computed empirically with intervals from the same bootstrap. No hypothesis tests of trend are performed.

Model changes, prompt version changes, fix deployments and documented incidents are marked on the weekly series. Model and prompt changes are dated from the run store itself, as the first window day on which a new model identifier or prompt version appears on terminal records. Code changes committed during the window that affect model selection are marked at their commit date and labeled as commit dates, because deployment dates are not recorded (Multimedia Appendix 1). Where at least 2 weeks of data lie on each side of a change, estimates before and after the change are reported with their intervals.

Two prespecified comparisons, each with an expected direction, are examined by estimation rather than testing. First, retry share (E3a) is expected to be higher in the deep reasoning class than in the fast triage class, reported as a difference with its 95% CI. Second, the number of resolved executions per completed intention (E2 divided by E1) is expected to be higher in the deep reasoning class, reported as a difference with its 95% CI. The expected directions were carried over from the initial plan, which drew on pre-protocol inspection of run-store data from before the window. Those data are not reported here and do not enter any analysis.

Five sensitivity analyses are prespecified: (1) the lower and upper bounds of rule D3; (2) intention grouping windows of 1 hour and 7 days (rule D4); (3) replacement of recorded zero costs on done records by an approximation computed from recorded token counts and the code rate table, which excludes cache tokens and is reported as an approximation; (4) imputation of cost for error records written without usage, using the median cost of error records with usage for the same agent and week; and (5) exclusion of weeks with documented incidents affecting the run store.

Extraction produces aggregates only. The queries are version-controlled, fixed to the window dates rather than to the time of execution, and run by SY at a recorded commit, and each extract file is hashed at creation. Gate outcomes are computed by the queries against the registered thresholds, which leave no discretion. No record-level rows, free text, encounter identifiers or input hashes leave the production environment.

### Data-Quality Gates and Failure Conditions

Each estimator depends on properties of the instrumentation that may not hold. The plan therefore specifies 4 data-quality gates (Table 3), each evaluated per stratum before any estimate is reported. A failed gate changes how an estimate is reported and is itself a reported result. If gate G1 or gate G3 fails at the level of the whole pipeline, the results paper will report the framework, the gate outcomes and the reasons for failure, and no pipeline-level cost estimates. The thresholds are conventional choices rather than derived values, and they are fixed before extraction. The share of done records with a recorded cost of 0 is reported for every stratum regardless of the gates, because it follows from per-call rounding rather than from a defect.

**Table 3.** Prespecified data-quality gates and the action taken when a gate fails.

| Gate | Check | Prespecified threshold | Action if the gate fails |
|---|---|---|---|
| G1 Terminal-state completeness | Executions with no terminal record 24 hours after start, after rule D1 | No more than 2% per stratum | Report E1 to E5 as bounds, computed with all such executions assigned first to error and then to completion; cost is 0 in both, because the records carry no usage |
| G2 Recording classification | Intention-key groups that are unclassifiable under rule D3 | No more than 2% of groups per stratum | Report all intention-level estimators as bounds |
| G3 Cost completeness | Terminal records with a recorded cost, and error records written without usage | At least 98% of terminal records with a recorded cost; error records without usage no more than 5% of resolved executions per stratum | First part: report cost estimators for the stratum as unmeasured. Second part: report cost estimators as lower bounds, with sensitivity analysis 4 |
| G4 Version coverage | Terminal records carrying both a model identifier and a prompt version | At least 98% per task class | Do not compute E7 for the task class |

### Instrumentation Status Before the Window

Each property that an estimator requires was assessed against the code at commit 687f45e295 and a version-locked extraction of pre-window records (Table 4), and each shortfall is mapped to the rule or gate that governs it. The extraction read only records that started before September 20, 2026, 23:00 UTC, and was run on November 19, 2026, by an AI coding assistant (Claude Opus 5.5) in a session operated by SY. The run store records every field that estimators E1 to E5 and E7 require on terminal records. Four properties on which their validity depends are not as the plan initially assumed. Every run is written as 2 records, so record counts are about twice execution counts. Terminal records are sometimes missing, and no process resolves them. Error records written on the exception path carry no usage. Recorded cost is rounded per call, in GBP pence. No entry for these properties exists in the deployment’s hazard log, so the source cited for each is the code at the pinned commit and the pre-window extraction.

**Table 4.** Status of the instrumentation before the measurement window, and the rule or gate that governs each property.

| Property required | Status before the window | Source | Consequence for the estimators | Rule or gate |
|---|---|---|---|---|
| Per-run recording of model, tokens, latency, cost, prompt version, input hash and status | Done records: all fields 100% populated. Error records: model, input hash, prompt version and finish time 100%; tokens, latency and cost 95.5%.<br>Non-terminal records carry no usage by design | Pre-window extraction (13,250 records) | Supports E1 to E5 and E7 on terminal records | G3, G4 |
| One record per execution | Every execution writes a non-terminal record and a separate terminal record; 6537 of 6604 key groups were exactly 1 of each | Write path at commit 687f45e295; pre-window extraction | Record counts overstate executions about 2-fold unless paired rows are removed | D1, G2 |
| Uniqueness of executions | Unique index on encounter, agent and input hash for done records only; no key had 2 done records | Schema at commit 687f45e295; pre-window extraction | Non-terminal and error records are unconstrained | D1 to D3, G2 |
| Terminal state written for every execution | 31 key groups held a non-terminal record with no terminal record; no process resolves non-terminal records; no fix at the pinned commit | Pre-window extraction; code | Unpaired records bias resolved-execution denominators | G1 |
| Usage retained on the exception path | Error records from exceptions carry no tokens, cost or latency (4.5% of error records) | Code; pre-window extraction | Cost of failed executions is understated | G3, sensitivity analysis 4 |
| Unit and provenance of recorded cost | GBP pence; provider-returned cost converted at 79 pence per US dollar, rounded per call; code rate table otherwise, with zero for unlisted models; 31.3% of done records recorded | Code; pre-window extraction | Cost of low-cost runs is understated or recorded as 0; token counts cannot reproduce cost | G3, sensitivity analysis 3 |
| Linkage of retries | No attempt counter, parent-run identifier or job identifier; failover usage not recorded; schema retry summed into 1 record | Code; schema | Only retries with unchanged input are linked; failover cost is not observed | D4, limitation |
| Model identifier | Non-terminal records carry the requested model and terminal records the model that answered | Code; pre-window extraction | Model mix is computed from terminal records | G4 |
| Separation of test runs | No test, sandbox or environment field in the run store | Schema at the pinned commit | Test runs in production are included | Limitation |

### Planned Reporting

The results paper will report the estimators by stratum in the format of Table 5, which is fixed here so that the strata, columns and suppression rules cannot change after extraction. Its primary figure will be the weekly series of cost per resolved execution, cost per completed intention and intention failure rate by task class over the window, with model and prompt changes, fix deployments and documented incidents marked, suppressed cells shown as gaps, and bootstrap intervals shown as shaded bands. Costs will be reported in GBP pence, with the fixed conversion rate stated so that readers can convert to US dollars.

**Table 5.**
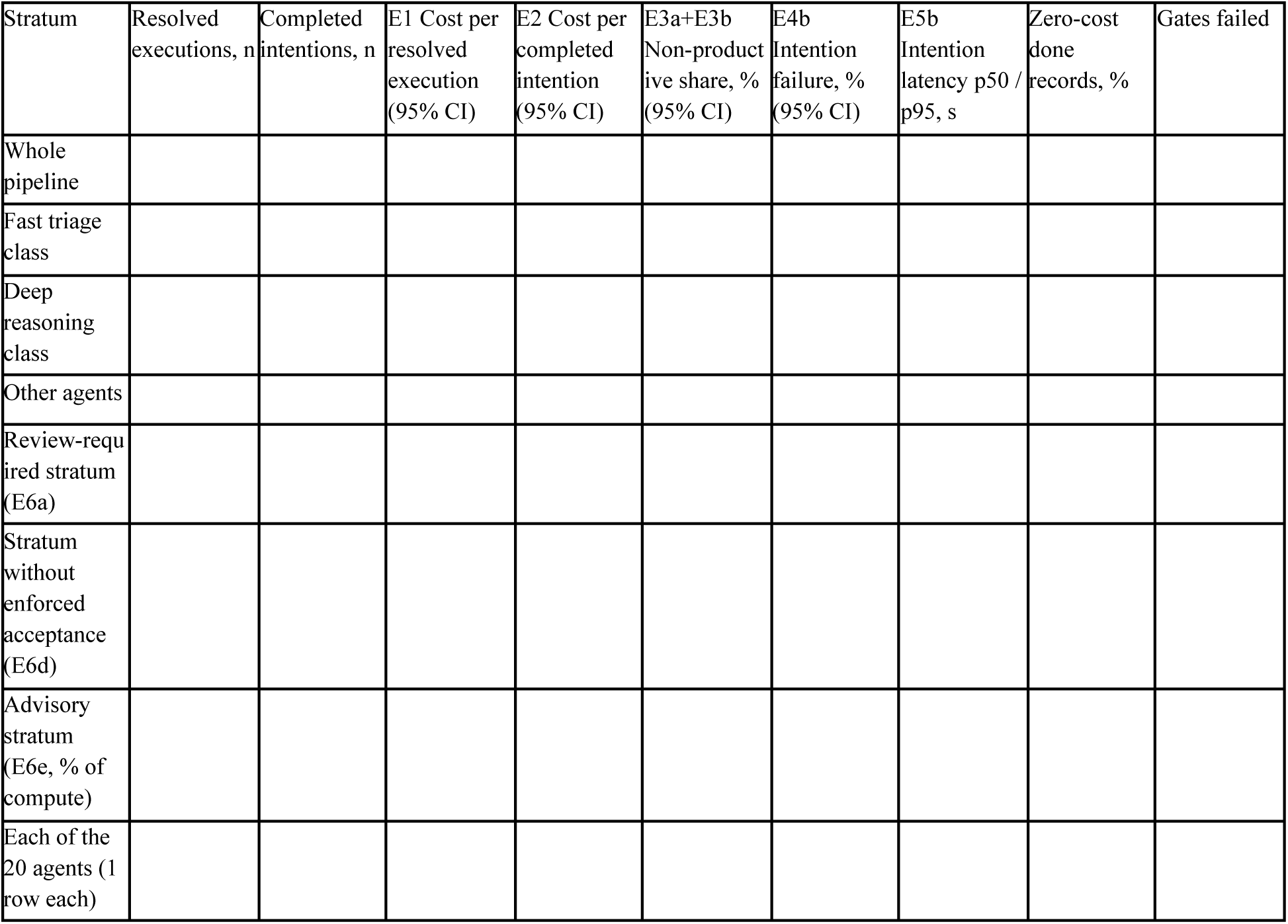
Prespecified reporting table for the results paper. No values are reported in this protocol. Costs are in GBP pence per unit, and intervals are 95% bootstrap percentile intervals over weeks. An estimator with a denominator below 50 in a stratum is reported as unmeasured.

### Scope and Exclusions

The framework measures compute resource use only. It contains no revenue, pricing, margin or staffing inference, reports unit costs and proportions rather than total expenditure, and supports no comparison of agent work with human work, which would require time-and-motion measurement of the human alternative. Engineering effort, infrastructure, oversight time and review time are real costs that the framework does not measure, so its estimates are not estimates of total cost of ownership. Agents outside the orchestrated pipeline are excluded because their compute is not recorded per unit of work.

### Registration and Elaborations to the Plan

The analysis plan was first written on September 21, 2026, and elaborated with the intention key and grouping window, the treatment of zero-cost terminal records, disjoint retry and failed-intention shares, the data-quality gates and their thresholds, the minimum-denominator threshold, the bootstrap method, the stratum for agents without enforced acceptance, the within-agent frontier and the sensitivity analyses. On November 19, 2026, the plan was revised again after the code and a pre-window extraction were checked: the recording rules were redefined on the 2-record write path, the cost unit was corrected to GBP pence with its rounding stated, the scope was restricted to the orchestrated pipeline, the test-run exclusion was removed because no flag exists, E6b, E6c and E6f were dropped, and 4 gates that no longer applied were removed. Every change is listed with its reason in the change log in Multimedia Appendix 1.

The complete plan and change log will be registered on Open Science Framework Registries, with a public time stamp, before any window data are extracted for this study. Any deviation from the registered plan will be listed with its reason in the results paper.

### Ethical Considerations

The protocol was developed from source code and configuration at a pinned commit, the action inventory and hazard log, and an aggregate extraction of pre-window run-store records. The planned analysis uses aggregate counts and sums of machine run telemetry from the run store: agent and model identifiers, token counts, cost, latency, status, prompt version and timestamps. No human participant is recruited, observed or surveyed. No identifiable personal data are analyzed: the encounter identifier, the input hash and the free-text error field are not extracted, no staff-level or approval data are used, and only aggregates leave the production environment. The authors therefore did not seek research ethics committee review, and no committee reviewed this classification. No participants are compensated.

## Results

The measurement window opened on September 21, 2026, and closes on December 13, 2026. Telemetry for the window is being recorded by the production system in routine operation, so data collection is under way and the study is declared as data existing. No window data had been extracted for this study when this protocol was submitted.

One exception is disclosed. On November 19, 2026, while companion manuscripts were being checked against the production database, aggregate counts were extracted that span the whole run store from May 2026 to November 19, 2026, and therefore include window records to that date. They were the number of records by status and by model identifier, the number of records by agent and status for 17 agents, including counts over the preceding 30 days for 10 of them, and the status composition of records sharing an intention key. No cost, token or latency value from the window was extracted, and no count was restricted to the window or broken down by week. The counts were extracted by an AI coding assistant in a session operated by SY, before the revision of the plan described above, and will not enter any analysis.

Neither a fix for the 2-record write path nor a process that resolves non-terminal records existed at commit 687f45e295 on November 19, 2026. Extraction will begin once the 24-hour terminal-state follow-up for the last executions in the window has ended, so no earlier than December 15, 2026, and after the plan is registered. The results are expected to be submitted for publication in 2027.

## Discussion

### Expected Findings

This protocol specifies estimators that link compute cost, latency, errors and retries to completed intentions in the orchestrated clinical pipeline of a multi-agent LLM system deployed in a single private outpatient clinic. Because the properties on which their validity depends are not all in place, with every run written as 2 records, terminal records sometimes missing and recorded cost rounded per call, the plan specifies in advance whether each estimate is reported as a point estimate, as a bound or as unmeasured.

Three kinds of result are expected. The first is the pair of prespecified comparisons, with a higher retry share and more resolved executions per completed intention in the deep reasoning class than in the fast triage class. The second is the gate outcomes, which are findings in their own right. Because no process resolves missing terminal records, gate G1 may fail for some strata, and estimates for those strata would then be reported as bounds. The third concerns the recorded cost itself. If the share of zero-cost done records seen before the window persists, cost estimators for the lowest-cost agents will understate their compute, and the token-based approximation will show by how much. Each of these outcomes will be reported.

Checking the plan against the code led us to revise a premise of the earlier plan. That plan treated a group of records sharing an input hash as the attempts behind 1 retried intention, and later as a mixture of retries and occasional duplicate writes. The write path shows that the second record is the rule rather than a defect: every execution is written twice. Retry detection from input hashes therefore requires a pairing step before any grouping, and the same consideration applies to any deployment that writes a pending record and a separate completion record for each run. We also found that the cost field, named in cents, holds pence. A unit cost read from such a field without checking the code would be wrong by the exchange rate.

### Comparison With Prior Work

Economic evidence on deployed clinical LLM tools concerns ambient documentation and is reported at the level of the clinician, the encounter or the subscription [10,11]. Randomized and multisite studies measured time and well-being rather than cost [1,2,4], and the editorial accompanying the trials concluded that the productivity case was not yet established [3]. The planned NHS evaluation will model cost consequences at trust level [12]. The present study measures a different quantity, the compute consumed per unit of completed work, which a subscription price does not reveal. It is complementary to these evaluations and could supply some of the resource-use measurements that a CHEERS-AI-conformant evaluation requires [27].

Benchmark studies report cost per query or per task under controlled conditions [13–15], and analyses of agent benchmarks have argued for joint reporting of cost and accuracy [16]. These designs do not include retries in production, failed intentions or fan-out across agents. Cost-of-pass, defined as the expected cost of producing 1 correct output, divides cost per attempt by the success rate [33]. Estimator E2 has the same structure. It applies it to production intentions rather than to benchmark items, and it adds the pairing of recording rows, the bounds for unclassifiable groups, the separation of agents without enforced acceptance and the data-quality gates that production telemetry requires. Estimator E7 places cost and latency on the same axes for the configurations of each agent, in the spirit of the joint cost-accuracy evaluation proposed for agents [16].

Of the 2 preprints closest to production, one reports completion after retries absorbed transient model errors [17], a rate that does not show the compute spent on the failed attempts, which estimators E3a and E3b report. The other reports a cost per appointment relative to manual scheduling [18], a comparison the present study does not make because it does not measure the human alternative. Production evidence that drafting tools can lengthen message turnaround [19] supports treating intention latency (E5b) as an outcome in its own right.

Prices for a fixed level of capability have fallen quickly and unevenly across tasks [20], while the price of running frontier models has risen with model size and reasoning demands [21]. A unit cost measured in one window is therefore bound to its model mix, prompt versions, price schedule and exchange-rate convention, and the frontier estimator is designed to be recomputed whenever a configuration changes rather than extrapolated. The same telemetry could serve the deployer monitoring and logging described in the Introduction [22–24]. The high-risk obligations of the European Union Artificial Intelligence Act for artificial intelligence in medical devices now apply from August 2, 2028 [34]. The cost of a draft offered for review (E6a) says nothing about whether the draft was used. The AI Airlock finding that oversight may weaken as users come to trust accurate outputs [25] is a reason to read any fall in that cost together with a measure of review fidelity, such as the one proposed in a companion manuscript [35], before it is interpreted as an efficiency gain.

### Limitations

This study has limitations. First, the usefulness of the estimators cannot be judged until the window closes. If gate G1 or gate G3 fails at the level of the whole pipeline, the results paper will report gate outcomes rather than cost estimates. Second, the setting is a single private outpatient clinic, and the study covers only the 20 orchestrated agents of a larger fleet; the messaging, voice and operations agents are excluded because their compute is not recorded per unit of work. The definitions are intended to be site-independent, but they have been checked against 1 schema only, and the estimates will not generalize to larger or differently organized services. Third, compute cost is not total cost of ownership. Engineering, infrastructure, oversight and review time are excluded, and compute unit costs read as total costs would favor automation. The estimates are intended to inform model routing, retry policy, prompt changes and monitoring, not adoption decisions. Fourth, the recorded cost is an imperfect measure of compute. It is converted at a fixed exchange rate, rounded to a whole penny per call so that many low-cost runs record 0, omits the usage of failover attempts within a call and of error records written on the exception path, and cannot be reproduced from stored token counts because cache tokens are not stored. The sensitivity analyses bound some of these effects but cannot remove them. Fifth, retries are linked only when they reuse the same input, because the run store has no attempt counter or parent run, which biases retry share downward and intention failure upward. Sixth, the run store has no field that marks a test run, so staff testing in production is included. Seventh, the plan was elaborated after the window opened, the expected directions of the 2 comparisons came from pre-protocol data, and aggregate whole-table counts that include window records were extracted for companion manuscripts before the final revision. This creates a risk of analytic flexibility, which is mitigated by listing every elaboration with its reason, disclosing the exception in full and registering the plan with a public time stamp before extraction. Eighth, this is a description and evaluation plan written by the people who built and operate the system: OE and SY hold equity in its developer, and SY will run the extraction. The gates and thresholds were fixed before extraction, and aggregate extracts with their generating queries will be available to editors and reviewers. Ninth, estimates depend on provider prices and the model mix during the window, both of which change quickly [20,21], so they will describe a configuration at a time rather than a stable property of the system.

### Dissemination

The results will be submitted for peer-reviewed publication as a results paper linked to this protocol by its International Registered Report Identifier, whatever the gate outcomes, including when gate G1 or gate G3 fails at the level of the whole pipeline. The aggregate extracts and their generating queries will be shared as described under Data Availability.

## Conclusions

The compute cost of completed clinical agent work can be specified as a set of estimators over run-level telemetry of the kind this deployment records, provided paired recording rows are separated from retries, missing terminal states are bounded and the unit and rounding of recorded cost are known. In this deployment, those conditions were not all met before the window opened, and the plan treats them as rules and gates rather than assuming them. Whether the estimators yield informative unit costs will be determined by the prespecified window, and replication in other deployments would be needed before the measures could be proposed as a reporting standard for clinical artificial intelligence.

## Supporting information

Multimedia Appendix 1 - agent map, specifications and change log

Multimedia Appendix 2 - TRIPOD-LLM checklist

## Acknowledgments

The authors thank the clinical and engineering teams at Wai Health Inc whose operating records make this work possible.

## Data Availability

The estimator definitions, recording rules, data-quality gates and analysis plan are given in full in this manuscript and in Multimedia Appendix 1. The weekly aggregate extracts from the prespecified window, with their version-controlled generating queries, will be available from the corresponding author under a data-sharing agreement after publication of the results paper, subject to suppression of cells with fewer than 5 events. No record-level, free-text or person-identifiable data will be shared. Instrumentation is described at module level, and the authors will facilitate verification of the deployed system by editors and reviewers under confidentiality on request. No proprietary source code is disclosed.

## Authors’ Contributions

All 3 authors contributed to the conceptualization of the study and to drafting and critical review of the manuscript, approved the final version and accept accountability for all aspects of the work. In addition, OE contributed to methodology and writing of the original draft; SY contributed to methodology, software (design and implementation of the telemetry instrumentation) and validation; and JD provided independent clinical safety review.

## Conflicts of Interest

OE and SY hold equity in Wai Health Inc, which developed and operates the system described. SY is Chief Technology Officer of Wai Health Inc. JD declares no competing interests.

Generative AI Disclosure The authors used large language model tools, including Claude (Anthropic), for drafting, editing and reference checking. On November 19, 2026, under the authors’ direction, Claude (Anthropic; Claude Opus 5.5) was used to check the manuscript against the system’s code and database and to revise it; the read-only database extractions cited in this protocol were run by Claude Opus 5.5 in a session operated by SY. The software described was itself developed with AI coding assistants, mainly Claude Code (Anthropic) and Cursor. All content was reviewed by the authors, who take full responsibility for the content of the manuscript. The generative AI conversations used in preparing the manuscript are provided in Multimedia Appendix 3.

Multimedia Appendix 1 Agent map, estimator and gate specifications, change points committed during the window, and change log of the analysis plan.

Multimedia Appendix 2 TRIPOD-LLM checklist with item locations, with items that do not apply to a protocol marked.

Multimedia Appendix 3 Generative AI conversations used in preparing the manuscript.

## Abbreviations

AI: artificial intelligence
CHEERS: Consolidated Health Economic Evaluation Reporting Standards
CHEERS-AI: CHEERS extension for interventions that use artificial intelligence
EHR: electronic health record
IRRID: International Registered Report Identifier
LLM: large language model
MHRA: Medicines and Healthcare products Regulatory Agency
NHS: National Health Service
TRIPOD-LLM: Transparent Reporting of a Multivariable Prediction Model for Individual Prognosis or Diagnosis, extension for large language models

