## Supplementary material for "What Does an Agent-Run Clinic Cost? Protocol for a Prospective Observational Study of the Compute Cost of Completed Work in a Clinical Agent Fleet": Multimedia Appendix 1 - agent map, specifications and change log

**Multimedia Appendix 1. Agent map, estimator and gate specifications, change points and change log**

**Part A. Agent map (frozen before extraction)**

Source: action inventory version 0.2 (September 8, 2026), accept column, and agent identifiers declared in the orchestrator's agent modules at commit 687f45e295. Each agent is mapped to its inventory row by the module path in the inventory's Path column.

**Agent identifier**

**Inventory row**

**Inventory accept class**

**Acceptance stratum**

**Task class**

clinical_summary

AA-01

Yes

Review required

Other

patient_summary

AA-02

Yes, two gates (sign then publish)

Review required

Other

action_points

AA-03

Partial

Acceptance not enforced

Fast triage

red_flag

AA-04

Advisory

Advisory

Fast triage

differential_dx

AA-05

Advisory

Advisory

Other

safety_netting_verifier

AA-06

Advisory

Advisory

Fast triage

drug_interaction

AA-07

Advisory

Advisory

Fast triage

coding_agent

AA-08

Advisory

Advisory

Other

guidelines_lookup

AA-09

Advisory

Advisory

Deep reasoning

investigation_suggester

AA-10

Advisory

Advisory

Deep reasoning

risk_score_recommender

AA-11

Advisory

Advisory

Other

mh_risk_triage

AA-12

Advisory, no accept step in code

Advisory

Fast triage

stopp_start_review

AA-13

Advisory

Advisory

Other

fit_to_fly_assessor

AA-14

Partial

Acceptance not enforced

Other

gp_letter

AA-15

Yes

Review required

Other

referral_letter

AA-16

Yes

Review required

Other

sick_note

AA-17

Yes, clinician signs

Review required

Other

letter_composer

AA-18

Yes

Review required

Other

note_composer

AA-19

Yes

Review required

Deep reasoning

note_critic

AA-20

Advisory, gates nothing

Advisory

Other

Totals: review required 7, acceptance not enforced 2, advisory 11; fast triage 5, deep reasoning 3, other 12.

Rules for agents not in the map: an agent identifier that appears in window records and is not listed above is assigned to the other task class, mapped to its inventory row by module path, and assigned to the acceptance stratum given by that row's accept class (Yes or Yes, weak: review required; Partial, No or any other wording: acceptance not enforced; Advisory or n/a: advisory). An identifier with no inventory row is reported as unmapped and contributes only to whole-pipeline estimates.

**Part B. Specifications**

All specifications operate on the run store, restricted to records whose start time lies in the window (September 20, 2026, 23:00 UTC to December 13, 2026, 24:00 UTC, which is September 21, 2026, 00:00 to December 13, 2026, 23:59 London time), with terminal records accepted if their start time is in the window and their finish time is no later than 24 hours after the window closes. Weeks are ISO weeks 39 to 50 of 2026 in London time. Fields used: agent identifier, model identifier, input hash, status, token counts, latency, cost, prompt version, start time, finish time. The encounter identifier, output hash, free-text error field and every other field are not read.

B1. Pairing (rule D1). Within each group of records sharing agent identifier, input hash and start time, order non-terminal records (status queued or running, with null token counts and null cost) and terminal records (status done, error, skipped or cost_capped) by record time, and pair them 1 to 1. Each paired non-terminal record is a recording row and is excluded. Each terminal record is an execution. Each unpaired non-terminal record is an execution with missing terminal state. A group in which a non-terminal record carries usage, or in which more non-terminal than terminal records share an identical start time with no way to order them, is unclassifiable (rule D3).

B2. Intentions (rules D2 and D4). Order executions with the same agent identifier and input hash by start time. The first execution opens an intention; every execution starting within 24 hours of the opening execution joins it; the first execution starting later opens a new intention. Sensitivity windows: 1 hour and 7 days.

B3. Costs. c is the recorded cost of a terminal record, in GBP pence; unpaired non-terminal records have c = 0. Error records with null token counts and null cost are counted as executions with c = 0 in the primary analysis and imputed in sensitivity analysis 4.

B4. Estimators E1 to E7 are computed exactly as defined in Table 2 of the manuscript, per stratum, for the whole window and per ISO week. Ratios of sums are computed by summing numerators and denominators over the units in the stratum; bootstrap replicates (2000) resample ISO weeks with replacement and recompute every ratio from resampled sums.

B5. Gates. G1: unpaired non-terminal executions divided by all executions, per stratum. G2: unclassifiable groups divided by all intention-key groups, per stratum. G3: terminal records with non-null cost divided by terminal records; and error records with null usage divided by resolved executions, per stratum. G4: terminal records with non-null model identifier and prompt version divided by terminal records, per task class. Thresholds and actions are in Table 3 of the manuscript.

B6. Sensitivity analysis 3. For done records with c = 0, an approximate cost is computed as input tokens multiplied by the input rate plus output tokens multiplied by the output rate of the code rate table in force at commit e636282a0e (July 14, 2026), in GBP pence per million tokens, for the model on the terminal record; models absent from the table remain 0. Cache tokens are not stored and are omitted. Results are labeled as an approximation.

B7. Outputs. Weekly and whole-window aggregates per stratum only, with cells below 5 events suppressed. No record-level output.

**Part C. Code changes committed during the window that affect model selection or retries**

Commit dates only; deployment dates are not recorded. They are marked on the weekly series and labeled as commit dates.

**Commit date**

**Commit**

**Change**

September 30, 2026

ed8ecf4546

Orchestrator gains an OpenRouter fallback chain (default deepseek/deepseek-v4-pro)

October 1, 2026

70c41da79c

A schema-invalid output is retried on a different model rather than the same one

October 2, 2026

490ebdef9f

Per-tenant processor, residency and zero-retention guard (inert unless the tenant configuration sets it)

October 18, 2026

2459021b2e

Orchestrator stops attempting Anthropic models, removing those candidates from every chain

October 23, 2026

9673ac215c

Model routing by benchmark added (inert, routing table empty)

No orchestrator agent's declared model or tier changed in the window; the last such change was committed on September 10, 2026.

**Part D. Change log of the analysis plan**

**Version and date**

**Change**

**Reason**

September 21, 2026

Plan first written

Not applicable

Versions to November 7, 2026

Intention key and 24-hour grouping window; zero-token terminal records; disjoint retry and failed-intention shares; 8 data-quality gates and thresholds; minimum denominator of 50; bootstrap over ISO weeks; unreviewed-effect stratum; within-agent frontier; sensitivity analyses

Methodological elaboration before extraction

November 19, 2026

Rule D1 redefined on the 2-record write path (pairing by agent, input hash and start time); call-log condition removed

Every run writes a non-terminal and a separate terminal record; the call-level log carries no run identifier and is not written by the orchestrator

November 19, 2026

Cost unit corrected from US cents to GBP pence; per-call rounding, fixed conversion rate and code rate table described; zero-cost share reported for every stratum

Cost is stored in pence, converted at 79 pence per US dollar and rounded per call

November 19, 2026

Sensitivity analysis replacing recorded cost by token-based recomputation changed to an approximation applied to zero-cost records; new sensitivity analysis imputing cost for error records without usage

Cache tokens are not stored, so recomputation cannot reproduce cost; exception-path error records carry no usage

November 19, 2026

Scope restricted to the 20 orchestrated agents; functional-domain strata (24) replaced by acceptance strata and agents

Only orchestrated agents write the run store; the inventory has 23 domains, and the inventory does not itemise the 29 actions without acceptance

November 19, 2026

E6b (review yield) and E6c (cost per committed effect) dropped; E6f (unlinked share), the fan-out allocation rule, the 7-day review follow-up, the approver-count rule and the third prespecified comparison removed; E6a redefined as cost per draft offered for review; E6d and E6e redefined on acceptance strata

E6b and E6c aggregate staff review decisions, which the study does not analyze; no proposal or approval record carries a run identifier

November 19, 2026

Gates G4 (call-log coverage), G5 (approval attribution), G6 (lineage completeness) and G7 (audit-trail integrity) removed; G8 (version coverage) renumbered G4; G3 redefined

The removed gates governed data sources no longer used

November 19, 2026

Exclusion of test and sandbox runs removed

No test, sandbox or environment field exists

November 19, 2026

Window fixed as ISO weeks 39 to 50 of 2026 (September 21 to December 13, 2026); daily aggregation for the final 28 days removed

Exact dates required before registration; the daily aggregation had no stated purpose

November 19, 2026

Extraction to be run by SY; gate outcomes computed by the queries

Named analyst required before registration
