## Supplementary material for "What Does an Agent-Run Clinic Cost? Protocol for a Prospective Observational Study of the Compute Cost of Completed Work in a Clinical Agent Fleet": Multimedia Appendix 2 - TRIPOD-LLM checklist

Checklist: TRIPOD-LLM (Gallifant et al, Nat Med 2025;31:60-69), Table 2, as published. Research design: LLM evaluation in a health care setting (H). LLM tasks of the agents studied: documentation generation, summarization and classification. The study evaluates the compute cost, latency and failure of deployed LLM agents, not the quality of their outputs, so items about output quality, annotation and performance are marked as not applicable. Results items are reported in the results paper.

**Section**

**Item**

**TRIPOD-LLM item (abridged)**

**Location in this protocol, or reason not applicable**

Title

1

Identify the study as developing, fine-tuning or evaluating an LLM, with task, population and outcome

Title

Abstract

2

TRIPOD-LLM for Abstracts

Abstract (Background, Objective, Methods, Results, Conclusions; registration line)

Background

3a

Health care context, use case and rationale, with references to existing approaches

Introduction, Background

Background

3b

Target population, intended use and intended users

Methods, Setting and System; Units of Analysis (acceptance strata)

Objectives

4

Study objectives and stage

Introduction, Objective

Data

5a

Sources of data

Methods, Data Sources

Data

5b

Relevant data points and their distribution

Methods, Data Sources; How a Run Is Recorded; Table 4 (pre-window population rates)

Data

5c

Dates of oldest and newest items

Methods, Prespecified Analysis Plan (window September 21 to December 13, 2026); pre-window extraction May 24 to September 20, 2026

Data

5d

Pre-processing and quality checking

Recording, Duplicate and Retry Rules; Data-Quality Gates (Table 3)

Data

5e

Handling of missing data and reasons for omitting data

Rules D1 to D3; gates G1 and G3; sensitivity analyses 3 and 4

Analytical methods

6a

LLM name, version and last date of training

Methods, Models (training dates not available to the authors; model mix measured from terminal records)

Analytical methods

6b

LLM development process

Not applicable: no model was developed or tuned

Analytical methods

6c

How text was generated, prompts and inference settings

Methods, Models (prompt version recorded per run); prompt text and inference settings are held in proprietary code and are available to editors and reviewers under confidentiality

Analytical methods

6d

Initial and post-processed output

Methods, Units of Analysis (drafts, findings and action points by stratum)

Analytical methods

6e

Classification thresholds

Not applicable: the study does not evaluate classification output

LLM output

7a

Metrics of output quality

Not applicable: output quality is not measured

LLM output

7b

Relevance of outcome metrics to the downstream task

Introduction (cost of completed work as the decision-relevant unit); Discussion, Comparison With Prior Work

LLM output

7c

Definition of outcome, calculation and date of inference

Estimators (Table 2); Prespecified Analysis Plan; Multimedia Appendix 1, Part B

LLM output

7d

Qualifications of assessors for subjective outcomes

Not applicable: no subjective assessment

LLM output

7e

Comparison with other LLMs, humans or benchmarks

Estimator E7 (configurations within agent); no comparison with human work (Scope and Exclusions)

Annotation

8a to 8c

Annotation process, annotators and their background

Not applicable: no annotation

Prompting

9a, 9b

Prompt design and data used to develop prompts

Not applicable: prompts are those of the deployed system and are not developed in this study; prompt versions are recorded and changes marked as change points

Summarization

10

Pre-processing before summarization

Not applicable: summarization quality is not evaluated

Instruction tuning or alignment

11

Instructions, data and evaluators

Not applicable

Compute

12

Compute or proxies required

The subject of the study: Methods, How a Run Is Recorded; Estimators

Ethical approval

13

Ethics committee approval or waiver

Methods, Ethical Considerations (no committee review sought, with rationale; no committee reviewed this classification)

Open science

14a

Funding and role of funders

Funding

Open science

14b

Conflicts of interest

Conflicts of Interest

Open science

14c

Where the protocol can be accessed

This manuscript; registration on Open Science Framework Registries before extraction

Open science

14d

Registration information

Abstract, Trial Registration line; Registration and Elaborations to the Plan

Open science

14e

Availability of study data

Data Availability

Open science

14f

Availability of code

Data Availability (extraction queries shared with the aggregates; system code not disclosed)

Public involvement

15

Patient and public involvement

Not reported in this protocol

Participants

16a to 16d

Flow and characteristics of data, number of events per analysis

Results paper; reporting format fixed in Table 5 and Figure 1

Performance

17

Performance against prespecified metrics

Results paper

LLM updating

18

Results of any LLM updating

Results paper; model and prompt changes marked as change points

Interpretation

19a

Overall interpretation

Discussion, Expected Findings (results paper for interpretation of results)

Limitations

19b

Limitations and biases

Discussion, Limitations

Usability in context

19c

Challenges in using the data

Instrumentation Status Before the Window (Table 4)

Usability in context

19d

Intended use, input, end user and level of autonomy

Setting and System; Units of Analysis (acceptance strata)

Usability in context

19e

Handling of poor-quality or unavailable input data

Not applicable: input data quality is not evaluated

Usability in context

19f

User interaction and expertise required

Units of Analysis (review-required drafts are accepted or signed by clinicians)

Usability in context

19g

Next steps for research

Discussion, Conclusions (replication in other deployments)
